# Manual vs. AI-Based Tractography: Assessing Fractional Anisotropy Consistency, Applicability and Methodological Implications

**DOI:** 10.1101/2025.04.25.25326403

**Authors:** Adrian Korbecki, Maja Gewald, Krzysztof Winiarczyk, Karol Zagórski, Agata Zdanowicz-Ratajczyk, Kamil Litwinowicz, Michał Sobański, Justyna Korbecka, Arkadiusz Kacała, Weronika Machaj, Anna Zimny

## Abstract

Tractography using diffusion tensor imaging (DTI) and constrained spherical deconvolution (CSD) provides valuable insights into the structure of white matter pathways. However, different methodologies may produce divergent fractional anisotropy (FA) values due to fundamental differences in their underlying approaches. This study compares FA measurements obtained using a manual DTI deterministic tractography method and an automatic AI-based approach via the TractSeg framework. Thirty healthy adults underwent brain MRI, and nine major white matter tracts were reconstructed using DTI-based vendor software and CSD with the AI-driven TractSeg.

FA measurements were analyzed using inter-rater reliability and agreement metrics, including intraclass correlation coefficients (ICCs). Results revealed substantial differences in FA between the two methods, with ICC values ranging from poor to moderate for most fibers. Normalization using FA values of the corpus callosum (CC) and comparison of relative values further highlighted impactful discrepancies for all of the fibers (p < 0.001). Manual DTI-based methods yielded higher FA values across most tracts, with the largest discrepancies observed in the CC and inferior fronto-occipital fasciculus. Conversely, AI-based TractSeg showed higher FA values for the uncinate fasciculus, demonstrating advantages for smaller, complex fibers.

Additionally, tract volume analysis showed that AI-based methods consistently produced larger tract volumes; however, volume differences did not align with FA ICC patterns. This indicates that volumetric discrepancies alone do not explain FA variability between methods. Despite high inter-rater reliability for manual measurements, significant inter-method differences indicate that FA values from the two methods are not interchangeable. Standardization is needed for reliable cross-study comparisons.

**Highlights:** Comparison of manual DTI and AI-based tractography in healthy adults.

Manual DTI yielded higher FA values; TractSeg favored smaller, complex tracts.

Tract volume differences did not explain FA variability across methods.

Poor-to-moderate FA agreement highlights need for cross-method standardization.

Study supports careful interpretation of tractography metrics across techniques.

## Introduction

Tractography is a technique that reconstructs white matter pathways from diffusion MRI (dMRI) data, offering insights into the brain’s structural connectivity (Le Bihan et al., 2001). This method includes both classic diffusion tensor imaging (DTI) approaches, which rely on principal eigenvectors, and advanced techniques such as constrained spherical deconvolution (CSD) (Auriat et al., 2015). By mapping individual white matter fibers, tractography provides valuable information about alterations in white matter integrity and the specific brain regions affected by disease. Importantly, it enables the investigation of microstructural changes beyond what conventional morphological assessments can reveal (Basser and Pierpaoli, n.d.; Jiang et al., 2017; Jones et al., 2013).

Advances in artificial intelligence (AI) have introduced automated, reader-independent segmentation methods that aim to overcome limitations associated with traditional approaches (Saha et al., 2020; Tchetchenian et al., 2023). While manual segmentation remains a gold standard due to its precision, it is labor-intensive, operator-dependent, and time-consuming. Automated AI-based methods, such as TractSeg, offer a promising solution by providing reliable, reproducible results with reduced user input, making them particularly attractive for large-scale studies (Wasserthal et al., 2019, 2018).

In this study, we compare fractional anisotropy (FA) measurements obtained from a manual DTI-based tractography approach performed by trained readers with results derived from the AI-based TractSeg framework employing CSD. The primary goal of this work is to determine whether FA values derived from these two methods can be directly or indirectly compared, focusing on potential differences and agreement levels between the approaches.

## Material and Methods

### Study population

This single-center observational study was conducted at the University Hospital of Wroclaw, Poland, between October 2020 and June 2021. The study cohort comprised 30 healthy, right-handed adults aged 26–74 years. Participants were selected based on strict inclusion and exclusion criteria to ensure homogeneity and the absence of health conditions that might influence the results.

Inclusion criteria required participants to be adults aged ≥18 years, in good physical and mental health, with no history of neurological or psychiatric disorders, systemic diseases, or substance abuse. Participants also needed to have no contraindications to MRI and were required to provide written informed consent.

Exclusion criteria included any neurological disorders affecting the central nervous system (CNS), severe psychiatric disorders, systemic diseases, use of psychoactive drugs or medications that could affect CNS function, alcohol or drug abuse, structural abnormalities detected by MRI, and contraindications to MRI.

Written informed consent was obtained from all participants. The study was approved by the local ethics committee (Komisja Bioetyczna, Wroclaw Medical University; approval No. KB-578/2020) and conducted in accordance with the Declaration of Helsinki and Good Clinical Practice guidelines.

### MRI image acquisition

All participants underwent brain MRI on a Philips Ingenia 3T system equipped with a 32-channel SENSE head coil. Scans were performed with standard patient positioning and coil setup to ensure consistent image acquisition across subjects. The imaging protocol included both a morphological T1-weighted 3D sequence and DTI, with the following parameters:

#### 1. 3D T1-weighted acquisition

T1-weighted images were acquired using a three-dimensional gradient echo sequence. The acquisition parameters were as follows: repetition time (TR) = 7.9 ms, echo time (TE) = 3.5 ms, flip angle = 8°, slice thickness = 1 mm, matrix size = 252 × 250, and field of view (FOV) = 250 mm. Images were acquired in the axial slice orientation to ensure optimal visualization of brain structures.

#### 2. DTI sequence acquisition

DTI was performed using an axial single-shot turbo spin-echo (TSE) echo-planar imaging (EPI) sequence with 12 diffusion encoding directions. The acquisition parameters included: b-value = 700 s/mm², TR = 3000 ms, TE = 59.3 ms, flip angle = 90°, matrix size = 140 × 140, FOV = 349 mm², voxel size = 2.5 mm², and slice thickness = 2.5 mm. Images were acquired in the sagittal slice orientation.

### White matter tract reconstruction

Fractional anisotropy (FA) was measured for entire white matter tracts, including the left and right corticospinal tracts (CST-left and CST-right), left and right arcuate fasciculus (AF-left and AF-right), left and right inferior fronto-occipital fasciculus (IFOF-left and IFOF-right), left and right uncinate fasciculus (UF-left and UF-right), and the corpus callosum (CC). These tracts were selected due to their precise definitions in the software, their relevance in both manual and automated assessments, and their critical contributions to motor, language, and cognitive functions.

Tract reconstruction was performed using two approaches: manually, with software provided by the MRI vendor, and automatically, with an AI-based technique implemented through the TractSeg framework.

#### 1. Manual tract reconstruction

Images were post-processed to correct for shear, eddy currents, and motion using the Diffusion Registration Tool (Philips Medical Systems). Deterministic DTI tractography was then performed with FiberTrak (Philips Medical Systems), employing a multiple-region-of-interest (ROI) strategy. This deterministic eigenvector-based approach relies on the principal diffusion direction within each voxel to guide streamline propagation, making it well-suited for reconstructing tracts with predominant, coherent fiber orientations (Christidi et al., 2016).

ROIs were manually placed at key anatomical locations suggested by the software guide, consistent with established landmarks (Christidi et al., 2016; Wakana et al., 2007). Typically, two freehand inclusion ROIs and, if necessary, one exclusion ROI were used to delineate each tract – except for the corpus callosum, where only a single ROI was required in the mid-sagittal plane. ROIs were drawn based on individual anatomy as depicted on overlaid color-coded DWI and T1 images, aiming to minimize errors and ensure precise tract delineation.

All tract segmentations were independently performed by three readers with limited experience in interpreting neurological images (approximately one year of radiology experience), following training by an experienced neuroradiologist and under continuous supervision during the measurement process. A minimum FA threshold of 0.15, a maximum turning angle of 27°, and a minimum track length of 10 mm served as tract termination criteria. After reconstructing the entire tract volume, FiberTrak calculated both the average FA values across the whole fiber and the tract volumes (Fig. 1). The readers were blinded to the results throughout the process.

**Fig 1.**
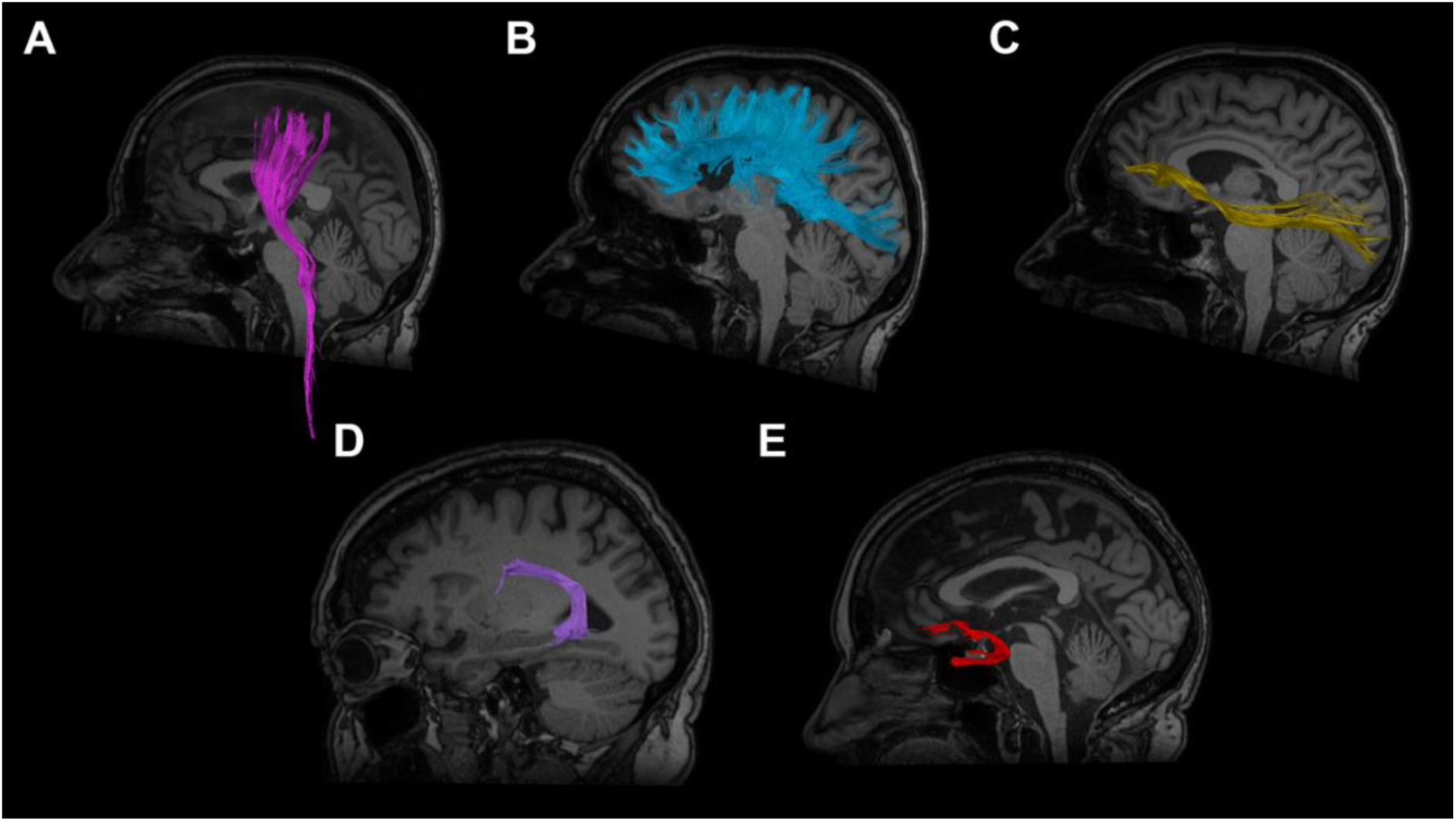
Reconstruction of the white matter tracts using FiberTrak (Philips Medical Systems) - left cortico-spinal tract (A), corpus callosum (B), left inferior fronto-occipital fasciculus (C), left arcuate fasciculus (D), left uncinate fasciculus (C) - overlaid on a high-resolution T1-3D sagittal slice

#### 2. AI-based method (TractSeg)

TractSeg is a novel method for direct white matter segmentation that utilizes an encoder– decoder convolutional neural network (CNN). In the initial steps, we employed a proprietary preprocessing pipeline provided by Hetalox Company. Diffusion-weighted images underwent preprocessing in MRtrix3, which included denoising, Gibbs artifact removal, eddy current correction, bias field correction, and skull stripping. Following this, rigid registration to MNI space was performed, and fiber orientation distributions (FODs) were estimated using constrained spherical deconvolution with the Dhollander algorithm for response function estimation (Dhollander et al., 2016; Tournier et al., 2019).

The resulting FODs were input into TractSeg, where a 2D encoder–decoder CNN generated tract probability maps in coronal, axial, and sagittal orientations. These maps were combined into a 3D representation for further refinement. The final output was a 72-channel image, with each channel representing voxel-wise probabilities for a specific tract (Fig. 2). For each tract, FA was automatically computed, and tract volumes were additionally estimated using TractSeg (Wasserthal et al., 2018). In our study, tracts were analyzed as whole structures without subdivision, allowing direct comparison of each reconstructed pathway. All processing was performed on a workstation running Ubuntu OS, equipped with a 16-core, 32-thread CPU processor and a standard GPU; in our study, the TractSeg pipeline was executed entirely on the CPU.

**Fig 2.**
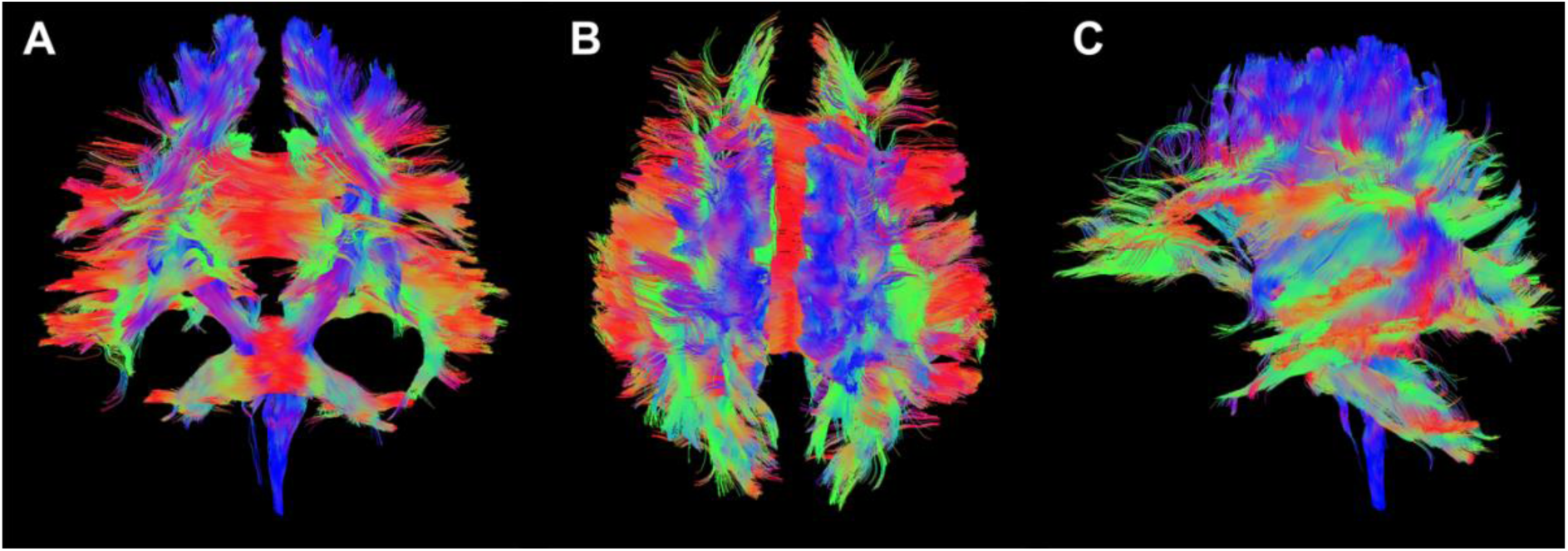
Reconstruction of the 72 white matter tracts using TractSeg with a CSD-based method, visualized in three planes: coronal plane (A), axial plane (B), and oblique plane (C). The color coding represents the orientation of the fibers: red indicates fibers running horizontally (left to right), blue indicates fibers running craniocaudally (up to down), and green represents fibers running anteroposteriorly (front to back).

### Relative FA values

For both tractography methods, relative FA values were calculated by normalizing each tract’s FA to the FA of the corpus callosum (CC) for each participant, serving as a reference structure. This normalization was applied to minimize variability between methods and enable a standardized, reliable comparison.

### Statistical analysis

To compare FA values and tract volumes obtained from manual DTI-based tractography and the AI-based algorithm, multiple statistical methods were used to evaluate agreement and reliability.

Inter-rater agreement was assessed for 30 participants using the intraclass correlation coefficient (ICC), employing a two-way mixed model with averaged measures for absolute agreement. Two ICC analyses were conducted: one among the three manual raters and another including the AI algorithm as a fourth rater. ICC values were interpreted using the following scale: < 0.5 as poor, 0.5–0.75 as moderate, 0.75–0.90 as good, and > 0.90 as excellent agreement (Koo and Li, 2016).

In the next step, ICC was used to assess agreement between mean FA values and tract volumes obtained with the manual DTI-based method and those calculated with TractSeg. Differences in relative FA values between the two methods were evaluated using the Related-Samples Wilcoxon Signed Rank Test, a nonparametric method for paired data. Statistical significance was set at p < 0.05. All analyses were performed using SPSS version 25.0 (IBM Corp., Armonk, NY, USA).

## Results

### Baseline characteristics

The study population included 30 healthy, right-handed participants, predominantly female (93,3 %) with a mean age of 54 ± 12.7 (range: 26-74). All participants were in good physical and mental health, with no history of neurological or psychiatric disorders, systemic illnesses, or substance abuse.

### Fiber reconstruction

The reconstruction of white matter tracts showed no significant differences between the methods. However, the number of fibers reconstructed using the DTI-based manual method differed considerably, particularly for the AF-right (Table 1).

**Table 1.** Number of reconstructed tracts: Manual vs AI-based methods.

| Tract | CST<br>- right | CST<br>- left | CC | IFOF<br>- right | IFOF<br>- left | AF<br>- right | AF<br>- left | UF<br>- right | UF<br>- left |
| --- | --- | --- | --- | --- | --- | --- | --- | --- | --- |
| Manual DTI-based method | 30 | 30 | 30 | 28 | 27 | 18 | 27 | 29 | 29 |
| AI-based TractSeg method | 30 | 30 | 30 | 30 | 30 | 30 | 30 | 30 | 30 |

Abbreviations: CST - cortico-spinal tract, CC - corpus callosum, IFOF - inferior fronto-occipital fasciculus, AF - arcuate fasciculus, UF - uncinate fasciculus

## Quantitative analysis

### DTI results - FA measurement

FA values were measured across the entire volume of selected white matter tracts using both the manual DTI-based deterministic method and the AI-based TractSeg method employing spherical deconvolution. Results, averaged across three readers, are summarized in Table 2.

**Table 2.**
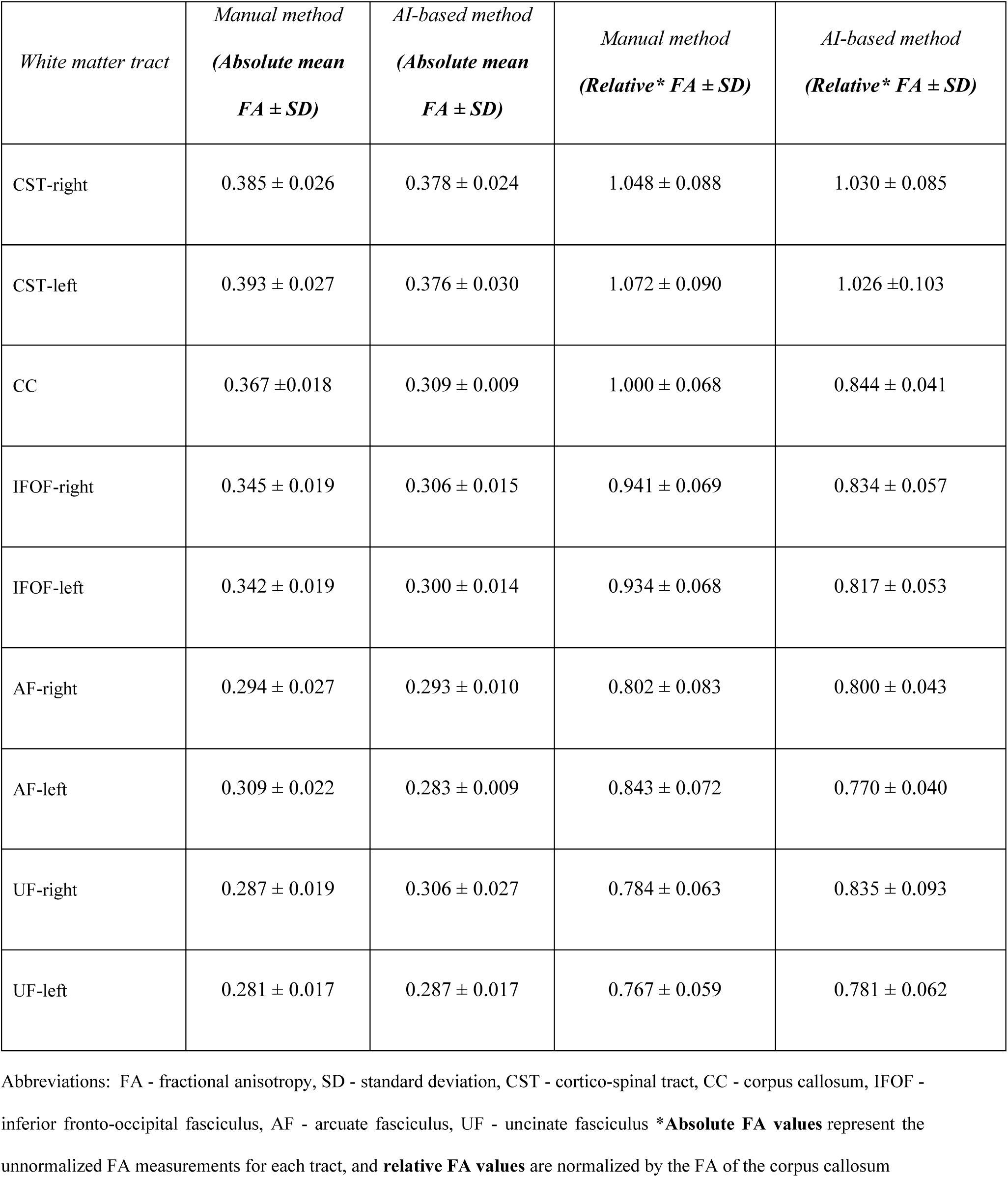
Absolute mean FA values and relative mean FA values for each white matter tract (Manual DTI-based vs AI-based method)

In general, FA values were slightly higher when measured using the manual method compared to the AI-based method for most white matter tracts. For example, in the corticospinal tract (CST), manual measurements yielded mean FA values of 0.385 ± 0.026 for the right CST and 0.393 ± 0.027 for the left CST, compared to 0.378 ± 0.024 and 0.376 ± 0.030, respectively, with the AI-based method. This pattern of slightly higher manual FA values was observed consistently across most tracts.

Interestingly, exceptions were found in the uncinate fasciculus (UF), where the AI-based method produced higher FA values. For the right UF, FA values were 0.287 ± 0.019 (manual) versus 0.306 ± 0.027 (AI), and for the left UF, they were 0.281 ± 0.017 (manual) versus 0.287 ± 0.017 (AI).

The second notable observation was that while differences in FA measurements between the two methods were generally small for most tracts. Substantial differences exceeding 10% were noted in specific fibers, particularly in the CC and the IFOF bilaterally. For the CC, the FA values were 0.367 (manual) versus 0.309 (AI), representing the largest discrepancy observed between the two methods. The IFOF also showed pronounced differences, as detailed in Table 2.

#### 1. ICC results – FA values

The inter-rater reliability among the three manual raters demonstrated excellent agreement for most white matter tracts, with ICC values ranging from 0.94 to 0.97 (Table 3). This indicates a high level of consistency in manual FA measurements across raters.

**Table 3.** Inter-rater reliability for FA measurements (ICC values for manual raters and manual vs AI)

| *White matter tract* | ICC (manual) | *ICC (manual and AI, as a fourth reader)* | ICC (manual vs AI) |
| --- | --- | --- | --- |
| CST-right | 0.96 | 0.91 | 0.75 |
| CST-left | 0.94 | 0.91 | 0.80 |
| CC | 0.86 | 0.78 | 0.53 |
| IFOF-right | 0.96 | 0.63 | 0.17 |
| IFOF-left | 0.94 | 0.62 | 0.17 |
| AF-right | 0.97 | 0.86 | 0.20 |
| AF-left | 0.95 | 0.79 | 0.32 |
| UF-right | 0.95 | 0.80 | 0.48 |
| UF-left | 0.95 | 0.88 | 0.66 |

Abbreviations: ICC - intraclass correlation coefficient, CST - cortico-spinal tract, CC - corpus callosum, IFOF - inferior fronto-occipital fasciculus, AF - arcuate fasciculus, UF - uncinate fasciculus

Inter-reader agreement, including the AI algorithm as a fourth reader, varied across tracts (Table 3). Excellent agreement (ICC = 0.91) was observed for the CST-left and CST-right. Good agreement (ICC = 0.78–0.88) was found for the CC, both AF, and UF. Moderate agreement (ICC = 0.62–0.63) was observed for the IFOF-left and IFOF-right.

When comparing the mean FA values obtained using the manual DTI-based method to those derived from the AI-based TractSeg method, the level of agreement varied across tracts and was generally low. Poor agreement was observed for the IFOF (both left and right), both AF, and the UF-right, with ICC values ranging from 0.17 to 0.48. Moderate agreement (ICC = 0.53–0.75) was noted for the CC, UF-left, and CST-right. Good agreement was found only for the CST-left, with an ICC value of 0.80, as detailed in Table 3.

#### 2. Comparison of manual and AI-based FA measurement - relative values

A detailed comparison of the relative FA values obtained from the manual and AI-based methods was performed, using the corpus callosum FA as a normalization factor for all other tracts in both methods. The analysis revealed no significant correlation between the relative values from the two methods (p < 0.001). Furthermore, statistical differences in the FA values for each white matter tract were identified, as shown in Table 4. The Wilcoxon Signed Rank Test confirmed significant differences (p < 0.001) across all tracts analyzed, indicating that the manual and AI-based methods produced consistently divergent results.

**Table 4.** Comparison between Manual and AI-based method for each white matter tract.

| *White matter tract* | *Manual method (Relative FA*<br>*± SD)* | *AI-based method (Relative*<br>*Absolute FA ± SD)* | *p-value* |
| --- | --- | --- | --- |
| CST-right | 1.048 ± 0.088 | 1.030 ± 0.085 | < 0.001 |
| CST-left | 1.072 ± 0.090 | 1.026 ± 0.103 | < 0.001 |
| CC | 1.000 ± 0.068 | 0.844 ± 0.041 | < 0.001 |
| IFOF-right | 0.941 ± 0.069 | 0.834 ± 0.057 | < 0.001 |
| IFOF-left | 0.934 ± 0.068 | 0.817 ± 0.053 | < 0.001 |
| AF-right | 0.802 ± 0.083 | 0.800 ± 0.043 | < 0.001 |
| AF-left | 0.843 ± 0.072 | 0.770 ± 0.040 | < 0.001 |
| UF-right | 0.784 ± 0.063 | 0.835 ± 0.093 | < 0.001 |
| UF-left | 0.767 ± 0.059 | 0.781 ± 0.062 | < 0.001 |

Abbreviations: FA - fractional anisotropy, SD - standard deviation, CST - cortico-spinal tract, CC - corpus callosum, IFOF - inferior fronto-occipital fasciculus, AF - arcuate fasciculus, UF - uncinate fasciculus

### DTI Results – volume measurement

Tract volumes were measured using both the manual DTI-based method and the AI-based TractSeg approach. The smallest tract volume observed with the manual method was for the AF-right (2560.01 ± 1902.19 mm³), while the smallest tract identified by the AI-based method was the UF-right (5980.76 ± 3444.13 mm³). In both methods, the largest reconstructed tract was the CC, with a mean volume of 95831.99 ± 22058.64 mm³ using the manual method and 314980.13 ± 32525.33 mm³ with the AI method (Table 5).

**Table 5.** Comparison of mean tract volumes (mm³) between manual DTI-based method and AI-based TractSeg method, with corresponding fold differences.

| White matter tract | Manual method (Mean volume in mm<sup>3</sup> ± SD) | AI method (Mean volume in mm<sup>3</sup> ± SD) | Fold Difference (AI / Manual) |
| --- | --- | --- | --- |
| CST-right | 9070.49 ± 4871.04 | 18240.48 ± 4490.74 | 2.01× |
| CST-left | 6898.09 ± 3914.46 | 21215.30 ± 5254.35 | 3.08× |
| CC | 95831.99 ± 22058.64 | 314980.13 ± 32525.33 | 3.29× |
| IFOF-right | 6164.94 ± 3697.12 | 35169.95 ± 6053.41 | 5.70× |
| IFOF-left | 5677.20 ± 4184.35 | 39998.89 ± 5841.94 | 7.05× |
| AF-right | 2560.01 ± 1902.19 | 47713.36 ± 8869.54 | 18.63× |
| AF-left | 4384.50 ± 2700.01 | 56330.31 ± 5681.79 | 12.85× |
| UF-right | 3451.01 ± 1594.45 | 5980.76 ± 3444.13 | 1.73× |
| UF-left | 2894.59 ± 1613.41 | 6927.93 ± 2835.89 | 2.39× |

Abbreviations: SD - standard deviation, CST - cortico-spinal tract, CC - corpus callosum, IFOF - inferior fronto-occipital fasciculus, AF - arcuate fasciculus, UF - uncinate fasciculus

Across all analyzed white matter tracts, the volumes obtained using the manual method were consistently lower than those produced by the AI-based approach. The extent of this difference varied substantially, with fold increases in AI-based volumes ranging from 1.73× for the UF-right to as high as 18.63× for the AF-right. Notably, this variation was not systematically associated with the anatomical size of the tract – larger tracts did not consistently exhibit smaller differences, nor did smaller tracts uniformly show greater discrepancies between methods. Full volume comparisons, including fold differences, are presented in Table 5.

### ICC – volume measurement

To evaluate the reliability of tract volume measurements, ICCs were calculated. When assessing agreement among the three manual raters using the DTI-based method, excellent inter-rater reliability was observed for the UF-right (ICC = 0.93) and UF-left (ICC = 0.92). Good agreement was found for the IFOF-left (ICC = 0.86), AF-left (ICC = 0.86), and IFOF-right (ICC = 0.81). Moderate agreement was seen for the AF-right (ICC = 0.65) and CC (ICC = 0.66). In contrast, the CST-left (ICC = 0.28) and CST-right (ICC = 0.36) showed poor inter-rater reliability, indicating greatest variability in volume measurements for these tracts among raters.

When the AI-based TractSeg method was included as a fourth rater, the overall agreement declined. Only the CC reached moderate agreement (ICC = 0.52), while all other tracts showed poor agreement.

Finally, when comparing the average volumes from the three manual raters to those obtained with the AI-based method, all ICC values remained within the poor range, with the highest observed agreement for the CC (ICC = 0.46). All ICC values for tract volumes are presented in Table 6.

**Table 6.** ICC values with 95% confidence intervals for tract volume measurements: manual raters, manual with AI as fourth rater, and manual (average) vs AI comparison.

| White matter tract | ICC (95% CI) - manual | ICC (95% CI) - manual and AI, as a fourth reader | ICC (95% CI) - manual vs AI |
| --- | --- | --- | --- |
| CST-right | 0.36 (0.13–0.58) | 0.30 (0.10–0.54) | 0.26 (–0.16–0.60) |
| CST-left | 0.28 (0.05–0.52) | 0.13 (–0.04–0.38) | 0.13 (–0.29–0.51) |
| CC | 0.66 (0.47–0.80) | 0.52 (0.31–0.72) | 0.46 (0.06–0.73) |
| IFOF-right | 0.81 (0.68–0.90) | 0.36 (0.14–0.61) | 0.16 (–0.28–0.54) |
| IFOF-left | 0.86 (0.75–0.93) | 0.43 (0.21–0.67) | 0.26 (–0.18–0.62) |
| AF-right | 0.65 (0.38–0.84) | 0.15 (–0.07–0.50) | 0.08 (–0.44–0.55) |
| AF-left | 0.86 (0.76–0.93) | 0.45 (0.22–0.68) | 0.25 (–0.20–0.61) |
| UF-right | 0.93 (0.87–0.96) | 0.12 (–0.05–0.37) | –0.19 (–0.56–0.24) |
| UF-left | 0.92 (0.86–0.96) | 0.27 (0.06–0.52) | –0.06 (–0.46–0.36) |

Abbreviations: CI – confidence interval, SD - standard deviation, CST - cortico-spinal tract, CC - corpus callosum, IFOF - inferior fronto-occipital fasciculus, AF - arcuate fasciculus, UF - uncinate fasciculus

## Discussion

The primary aim of this study was to compare FA measurements obtained using the manual DTI-based deterministic method with those derived from the AI-based TractSeg framework employing CSD. By doing so, we sought to determine whether the results from these two methods could be directly or indirectly compared. Our study focused on a cohort of 30 healthy individuals without any neurological symptoms or morphological abnormalities to ensure that the findings were not influenced by pathological variability.

The main finding of this study is that FA values differ significantly between the two methods. Specifically, FA values obtained using the manual DTI-based method were generally higher across most white matter tracts compared to those derived from the AI-based TractSeg method. The largest discrepancies were observed in the CC and the IFOF bilaterally, where FA values from the manual method substantially exceeded those from the AI-based method (Ressel et al., 2018; Tallus et al., 2023). An exception to this trend was observed in the UF, where the AI-based method produced slightly higher FA values.

These findings were corroborated through ICC analysis, which was conducted using two different approaches. First, we treated the AI-based method as a fourth reader alongside the three manual readers. In this comparison, excellent agreement was observed for the CST-left and right, while good agreement was found for the CC, AF and UF bilaterally. Moderate agreement was found for the remaining fibers. Second, we compared the averaged manual measurements directly to those obtained from the AI-based method. In this direct comparison, the highest agreement observed was good and limited to the CST-left. For the remaining fibers, agreement was poor or moderate at best, highlighting a lack of consistency between the two methods. These findings demonstrate that the FA values derived from the manual and AI-based methods are fundamentally incomparable for direct application across studies (Calamuneri et al., 2018).

Furthermore, to mitigate these discrepancies, we attempted normalization by comparing relative FA values for each fiber against the FA of the CC within the method. While this approach reduced variability within each method, it did not resolve the inconsistencies between them. The relative values again highlighted significant divergence between the two methods, as evidenced by the Wilcoxon Signed Rank Test, which consistently identified significant differences (p < 0.001) across all tracts analyzed.

Additionally, when assessing tract volumes, we found that all manually reconstructed tracts using the DTI-based method yielded consistently smaller volumes compared to those generated with the AI-based TractSeg approach. This difference ranged from a 1.73-fold increase for the UF-right to an 18.63-fold increase for the AF-right in the AI-based method. At the same time, the agreement in volume measurements, as assessed by ICCs, followed a different pattern.

When treating the AI method as a fourth reader, only the CC showed moderate agreement (ICC = 0.52), while all other tracts demonstrated poor agreement. Similarly, when comparing the average of manual volumes to the AI-derived volumes, all tracts showed poor agreement, with the CC exhibiting the highest ICC (0.46).

These findings, in conjunction with a detailed analysis of FA ICCs, suggest that differences in tract volume do not account for variability in FA reliability. For instance, tracts with relatively small fold changes in volume (e.g., UF-right) or relatively high volume agreement (e.g., CC) did not necessarily show better FA ICCs. This lack of correspondence indicates that agreement in FA values does not reliably mirror agreement in volume – whether assessed through mean comparisons or ICCs.

Reconstructed volumes do not equate to actual anatomical volume and may be influenced by numerous methodological factors, including tract curvature, branching, and modeling assumptions. Diffusion MRI has inherently limited spatial specificity due to factors like voxel size, partial volume effects, and its inability to resolve fine microstructural details such as axonal packing and organization (Jones et al., 2013).

Therefore, the discrepancies in tract volume and ICCs are most likely attributable to methodological differences between deterministic DTI-based tractography and the AI-based CSD TractSeg approach. Importantly, we aimed to keep the tracking parameters as consistent as possible between both methods (e.g., FA threshold), thereby minimizing the likelihood that these settings contributed to the observed differences in volume or agreement metrics.

Taken together, these findings – encompassing both FA and volume measurements – indicate that FA measurements from manual DTI-based methods and AI-based TractSeg are not interchangeable, even after normalization. The observed discrepancies likely reflect fundamental differences in how these methods model diffusion, define fiber orientations, and delineate white matter pathways (Mormina et al., 2016; Ressel et al., 2018).

The use of relative values for normalization in this study raises important questions and potential challenges. Normalization inherently requires a reference structure, and we chose the CC as it is one of the most easily identifiable tracts in the ROI-based DTI manual method and demonstrates high repeatability between readers. However, this approach introduced issues, as the CC displayed the most substantial differences in FA values between the two methods - the manual DTI-based method yielded a mean FA value of 0.367 ± 0.018, while the AI-based TractSeg method produced a significantly lower value of 0.309 ± 0.009. This discrepancy suggests that using the CC as a normalization reference may be inappropriate.

The structural characteristics and methodological differences disproportionately influence CC FA values. The CC consists predominantly of parallel fibers, which may lead to higher FA values in the manual DTI-based deterministic approach (Tallus et al., 2023; Vos et al., 2012). This method relies on identifying a single principal eigenvector per voxel, which is well-suited for highly organized tracts like the CC. Conversely, the AI-based TractSeg method, which employs CSD to address crossing and kissing fibers, and relies on fiber orientation distribution function (fODF) incorporates more complex diffusion signals. As a result, TractSeg may produce lower FA values for such tracts (Calamuneri et al., 2018).

Future studies should consider alternative reference structures that exhibit greater consistency between methods. Potential candidates include tracts with smaller inter-method differences or setting the ROI within gray matter, which may provide a more stable normalization baseline. Notably, the higher FA values observed in the CC align with findings in the literature, further supporting the validity of our observations (Tallus et al., 2023). These findings underscore the need for careful selection of normalization references to ensure meaningful comparisons across methods.

In our study, we observed that FA values in the UF were higher when measured using the AI-based TractSeg method compared to the manual DTI-based method. Specifically, the AI-based method yielded mean FA values of 0.306 ± 0.027 for the UF-right and 0.287 ± 0.017 for the UF-left, whereas the manual method produced slightly lower values of 0.287 ± 0.019 for the UF-right and 0.281 ± 0.017 for the UF-left. This finding is noteworthy, as the UF was the only fiber where the AI-based method resulted in higher FA values.

We hypothesize that this discrepancy may be attributed to the anatomical characteristics of the UF and the methodological differences. The UF is a relatively small and anatomically complex fiber, which makes it challenging to delineate accurately using manual ROI in the DTI-based approach. In such cases, the reliance on a single principal eigenvector per voxel in the DTI method may limit the ability to fully capture the fiber’s diffusion properties, especially in regions with crossing fibers or partial volume effects (Auriat et al., 2015; Calamuneri et al., 2018; Mormina et al., 2016; Reijmer et al., 2012).

Conversely, the AI-based TractSeg method, unlike the DTI-based method, it is not constrained by an FA threshold that excludes voxels with less dominant diffusion directions. This allows it to identify smaller or more complex fibers, potentially explaining the higher FA values observed for the UF (Tournier et al., 2008). These results suggest that CSD-based approaches may be better suited for the analysis of smaller, intricate tracts, where conventional eigenvector-based methods may struggle to provide accurate measurements (Auriat et al., 2015; Calamuneri et al., 2018; Mormina et al., 2016).

One key difference between our study and literature is the choice of tract termination thresholds, particularly the minimum FA limit (Tallus et al., 2023). We used the FiberTrack default threshold of 0.15 for all fibers, while Tallus et al. applied a more stringent FA limit of 0.50 for the CC and 0.3 for other tracts. Other parameters, such as angular deviation and minimum tract length, were consistent between studies.

Using a uniform FA threshold ensured standardization across tracts in our study but may have influenced outcomes by including voxels with lower anisotropic diffusion, particularly in regions with complex geometries. The higher FA thresholds used by other researchers contribute to differences in tractography results between studies (Schilling et al., 2021).

Our study demonstrated exceptionally high ICC among the manual measurements performed by the three readers, despite their relatively limited radiological experience. This finding suggests that, with proper training and supervision of an experienced neuroradiologist, individuals can perform tractography measurements consistently and with high reliability (Danielian et al., 2010; Fox et al., 2012). The high agreement observed in our study underscores the importance of following standardized protocols for ROI placement, as outlined in the guidelines provided by the FiberTrack software.

This suggests that researchers and clinicians who are proficient in tractography can rely on manual measurements for comparative studies without significant concerns about inter-reader discrepancies. Moreover, it reinforces the utility of manual methods as a robust benchmark against which other approaches, can be evaluated (Ades-Aron et al., 2018).

Finally, the DTI-based manual approach, while thorough, posed significant challenges in our study. Despite following the instructions and repeated attempts by the readers, certain fibers could not be reliably tracked. For instance, the AF-right proved particularly problematic, even for supervising neuroradiologists. This highlights a critical drawback of the manual method: the inability to track certain fibers reliably, particularly smaller or more complex tracts, as the eigenvector-based method often struggles with partial volume effects and regions of complex fiber geometry. Additionally, this method is highly time-consuming, requiring multiple steps, including opening and preprocessing each examination, overlaying T1-weighted images, carefully selecting ROIs, and manually excluding incorrect fibers. This process demands a solid understanding of tract anatomy, as errors in ROI placement or misidentification of fibers could result in incomplete or inaccurate tractography, which alters the diffusion values (Maier-Hein et al., 2017; Rheault et al., 2020).

In contrast, the AI-based CSD TractSeg method demonstrated significant advantages in terms of efficiency, reliability, and automation (Faiyaz et al., 2023; Wasserthal et al., 2019; Zhang et al., 2020). The full TractSeg pipeline, which reconstructs all 72 white matter tracts per patient, was completed in approximately 30 minutes. However, since multiple patients (up to 32) could be processed simultaneously – one per CPU thread – the effective processing time per patient was less than one minute. This background processing capability allowed radiologists and researchers to continue other tasks concurrently. The method’s speed and scalability depend on the number of available CPU threads, and further acceleration is possible through CUDA-enabled GPU processing (Joshi et al., 2024).

The TractSeg method also offered superior consistency and objectivity, as the results were identical for the same patient across previously done repeated analyses, eliminating the variability introduced by human operators (Tchetchenian et al., 2023; Wasserthal et al., 2019). However, this method is not without its drawbacks, particularly its dependence on the preprocessing pipeline and the requirement for efficient hardware to optimize performance.

Our study has several limitations that should be considered when interpreting the results. First, the sample size was relatively small, comprising only 30 healthy individuals, which may limit the generalizability of our findings. Second, the study focused on a subset of nine selected white matter tracts, and it would be valuable to expand future analyses to include additional fibers for a more comprehensive assessment. Another limitation relates to the quality of the DTI examinations. The scans were acquired in sagittal plane to include the cervical spinal cord, which may have introduced challenges for both methods. Despite this, both the manual DTI-based and AI-based TractSeg approaches successfully tracked fibers in most cases, consistent with findings from prior studies, which also utilized DTI data with a limited number of vectors and lower b-values (Calamuneri et al., 2018; Tallus et al., 2023). Notably, the DTI-deterministic vendor software was unable to efficiently process higher-resolution diffusion data, which limited our ability to use more advanced acquisition protocols.

Additionally, the relatively limited experience of the manual raters could be considered a limitation. This was mitigated through initial training and supervision by an experienced neuroradiologists. Addressing these limitations in future research would further strengthen the validity and applicability of the findings.

## Conclusion

Our findings indicate that FA values obtained from the manual DTI-based method and the AI-based CSD TractSeg method cannot be directly or indirectly compared due to significant methodological differences. While we attempted normalization techniques, these approaches proved inadequate, underscoring the need for future research to explore proper normalization strategies or reference structures.

The AI-based CSD TractSeg approach demonstrates significant advantages. It offers a fast, reproducible, and objective method that reliably produces comparable results, even for complex and smaller tracts that pose challenges for the manual DTI-based approach. Additionally, the AI-based method is capable of generating 72 fibers at one analysis, far exceeding the capacity of manual methods, and does so with consistent precision and efficiency.

While the manual DTI-based method provides greater flexibility and remains a gold standard for certain fibers, it is time-intensive, operator-dependent, and less reliable for smaller or intricate tracts. Thus, the automatic, AI-based approaches are promising techniques for future research and clinical applications in white matter analysis.

## Data Availability

All data produced in the present study are available upon reasonable request to the authors.

